# Bayesian Shrinkage Priors in Zero-Inflated and Negative Binomial Regression models with Real World Data Applications of COVID-19 Vaccine, and RNA-Seq

**DOI:** 10.1101/2022.07.13.22277610

**Authors:** Arinjita Bhattacharyya, Riten Mitra, Shesh Rai, Subhadip Pal

## Abstract

**Background:** Count data regression modeling has received much attention in several science fields in which the Poisson, Negative binomial, and Zero-Inflated models are some of the primary regression techniques. Negative binomial regression is applied to modeling count variables, usually when they are over-dispersed. A Poisson distribution is also utilized for counting data where the mean is equal to the variance. This situation is often unrealistic since the distribution of counts will usually have a variance that is not equal to its mean. Modeling it as Poisson distributed leads to ignoring under- or overdispersion, depending on if the variance is smaller or larger than the mean. Also, situations with outcomes having a larger number of zeros such as RNASeq data require Zero-inflated models. Variable selection through shrinkage priors has been a popular method to address the curse of dimensionality and achieve the identification of significant variables.

**Methods:** We present a unified Bayesian hierarchical framework that implements and compares shrinkage priors in negative-binomial and zero-inflated negative binomial regression models. The key feature is the representation of the likelihood by a Polya-Gamma data augmentation, which admits a natural integration with a family of shrinkage priors. We specifically focus on the Horseshoe, Dirichlet Laplace, and Double Pareto priors. Extensive simulation studies address the efficiency of the model and mean square errors are reported. Further, the models are applied to data sets such as the Covid-19 vaccine, and Covid-19 RNA-Seq data among others.

**Results:** The models are robust enough to address variable selection, and MSE decreases as the sample size increases, having lower errors in *p > n* cases. The noteworthy results showed that the adverse events of Covid-19 vaccines were dependent on age, recovery, medical history, and prior vaccination with a remarkable reduction in MSE of the fitted values. No. of publications of Ph.D. students were dependent on the no. of children, and the no. of articles in the last three years.

**Conclusions:** The models are robust enough to conduct both variable selections and produce effective fit because of their high shrinkage property and applicability to a broad range of biometric and public health high dimensional problems.

## 1 Introduction

The extension of linear models to generalized linear models (GLMs) introduced by [1] framework to handle data that are not typically modeled using a normal distribution (e.g., binary, count data) is a moment of immense success in the history of statistics. Most of the real-life datasets have number of explanatory variables (*p*) greater than the number of observations (*n*).The methodology for analyzing RNA sequencing data is rapidly expanding. Methods having application of shrinkage, flexibility of the designs, are highly in demand [2]. High-dimensional predictor selection and sparse signal recovery are routine statistical and machine learning practices.Sparsity relies on the property of a few large signals among many (nearly) zero noisy observations. A common goal in high-dimensional inference is to recover the low-dimensional signals observed in noisy observations[3]. The idea of global-local shrinkage hierarchies[4] has become the foremost research areas in Bayesian literature that incorporates heavy tailed prior distributions for coefficients in generalized linear regression models. In the exponential rise in the development of research in shrinkage priors, works that have gained mass popularity are [5], [6], [7], [8], [9], [10], [11], [12], [13], [14] among many. An overview of several shrinkage priors with several data applications is given in [15]. Here we discuss the posterior simulation for negative binomial (NB) regression and Zero-inflated Negative binomial (ZINB) for count data. Our main focus is the utilization of the Polya-Gamma (PG) data augmentation strategy [16] which utilizes Polya-Gamma random variables to enhance posterior simulations. The performance of three different priors Horseshoe (H)[17],Dirichlet Laplace (DL) [14], Double Pareto (DP)[13] are measured and also the method is applied to benchmark data sets. Bayesian global-local (GL) shrinkage estimation is the state-of the art for Gaussian regression models, extension to non-standard regression techniques such as Poisson and NB are the ones that we concentrate in our methodology. Here two extensions of the global-local shrinkage framework is implemented. Firstly, the utilization of the PG data augmentation technique to generate simple algorithms for sampling with NB and ZINB regression likelihoods. Results show that the priors are highly competitive on the basis of mean square errors (MSE). Extensive simulation studies and real data applications are conducted to evaluate the performance of the these priors with respect to prediction accuracy and MSE for variable selection.

The rest of the section follows as section 2 describes in detail the tradition methods such as Poisson and NB and their Bayesian counterpart such as BNB and BZINB.Section3 explains the simulation details and parameters, followed by results 4. Real data scenarios are explained in Section 5, finally summarizing manuscript with a discussion 6.

## 2 Method

### 2.1 Poisson Regression

In modeling the number of times an event occurs a generalized linear model (GLM) such as Poisson or negative binomial regression is commonly applied.Let *y* = (*y*_1_, *y*_2_, …, *y*_*n*_) denote the vector of *n* count measurements of a dependent variable of interest, and *x*_*i*_ = (*x*_*i*,1_, …, *x*_*i,p*_) denote the vector of predictors (explanatory variables, covariates) associated with the response *y*_*i*_. Let *X* = (*x*_1_, …, *x*_*n*_) be the *n* × *p* matrix of explanatory variables. The GLM contains Poisson distribution which has a probability function 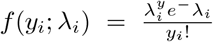 where the mean and variance are equal. *E*(*y*_*i*_) = *var*(*y*_*i*_) = *λ*_*i*_. The log-likelihood is given by 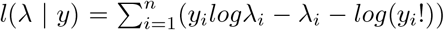. In GLM, a non-linear transformation or a link function of the mean response *λ*_*i*_ is applied which is a linear function of the covariates *X* [18],[19]. The link function for Poisson regression is 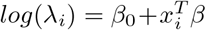, where *β*_0_ is the intercept and *β* is the vector regression coefficients. Poisson distribution depending on single parameter *λ* is equi-dispersed. If the Poisson mean is assumed to have a random intercept term and this term enters the conditional mean function in a multiplicative manner, the following relationship is achieved [20]:

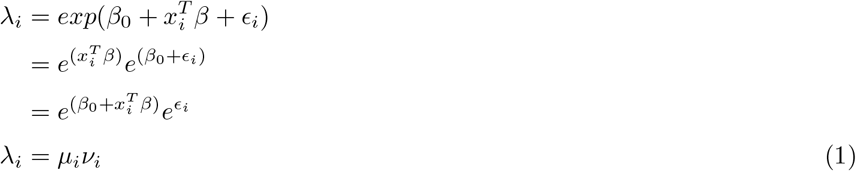

Here, *exp*(*β*_0_ + *ϵ*_*i*_) is defined as the random intercept; 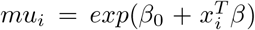is the log-link function between the mean *E*(*y*_*i*_) and the independent fixed covariate matrix *X*. The real data, however, often shows that the variance is larger than the mean, which is called overdispersion. The two-parameter negative binomial distribution has more flexible and can efficiently model overdispersed count data leading to correct standard errors and inferences [21]. Many parametric models for count data are obtained by additionally introducing a heterogeneity term in the Poisson model. Unobserved heterogeneity is usually included as a multiple of the Poisson mean. *y* | *µ, ν* ∼ *Poi*(*µν*) and the random heterogeneity term *ν* ≥ 0 is integrated out to obtain the distribution of *y* | *µ*. These model structures are well-known as doubly stochastic Poisson by Cox [22] and a Cox process by Kingman [23]. In general, *E*(*ν*) = 1 is the setting for several leading models. Different distributions of *v* leads to various generalizations of Poisson and here the Poisson-Gamma mixture is explained which leads to Negative Binomial distribution.

### 2.2 Negative Binomial as Poisson-Gamma Mixture

The NB model is derived from a Poisson-gamma mixture distribution [18]. The interpretation and derivation of NB from Poisson-Gamma is detailed in [24]. The heterogeneity parameter is assumed to have a Gamma distribution. *ν*_*i*_ ∼ *Gamma*(*d, b*). The two-parameter Gamma distribution is represented as

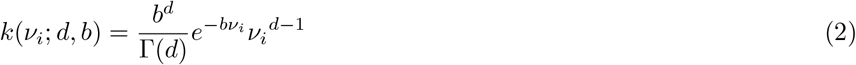

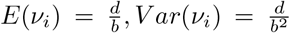, setting *E*(*ν*_*i*_) = 1 we get *d* = *b*, leading to a one-parameter Gamma distribution with 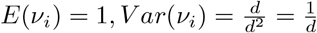. Now the Poisson model *Poi*(*µν*) can have easier interpretation if worked with the transformation *λ* = *μν*, i.e. 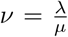. The Jacobian is obtained as 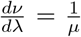. The probability density function (p.d.f) of *λ* is then given as

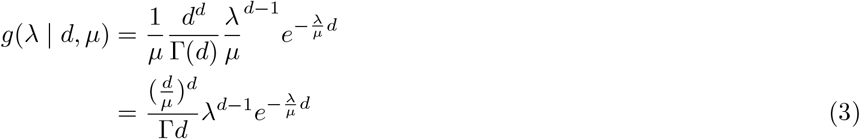

The Poisson-gamma mixture is

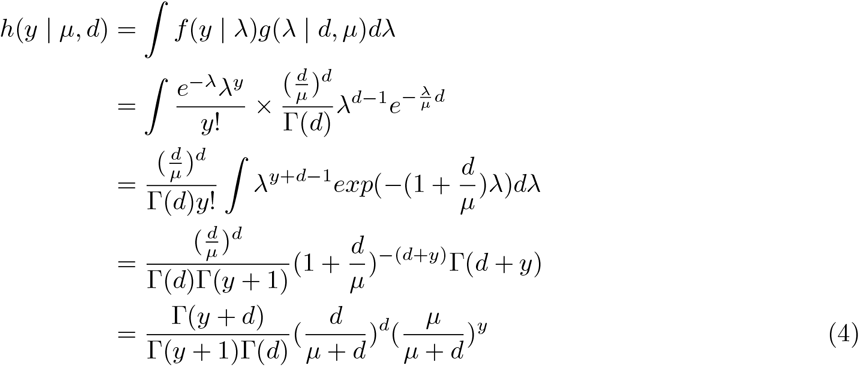

The property of the gamma function is utilized in getting the above form: 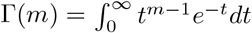 for any 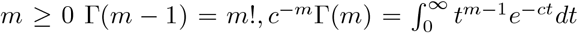 for any *c* ≤ 0.

The equation (4) can also be represented as

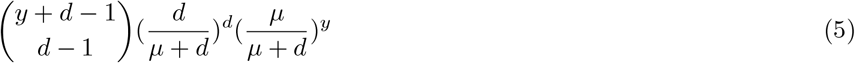

Taking 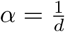we get the probability mass function (p.m.f) of the NB distribution as

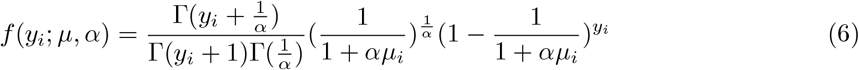

Here, *E*(*y* | *µ, α*) = *µ, V ar*(*y* | *µ, α*) = *µ*(1 + *αµ*) ≥ *µ*, since *α* ≥ 0. The variance can also be represented as 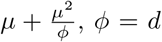 is the dispersion parameter. Most often it is expressed as 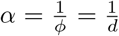, *α* is the parameter responsible for heterogeneity and models the over-dispersion amount in the data An alternative parametrization of NB used in many references as well as algorithms is

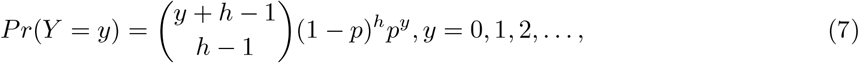

where 0 *< p <* 1 and *h* ≥ 0. Then 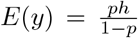and 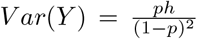 Letting 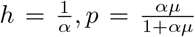, yields the same parametrization of NB distribution in (6) and *h* = *d* in (5).

The log-likelihood of NB can be expressed as

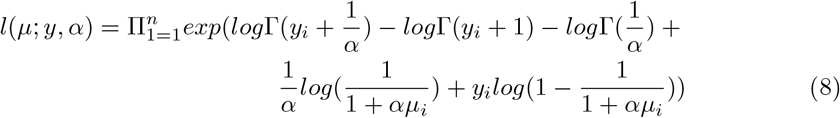

The mean of (7) is

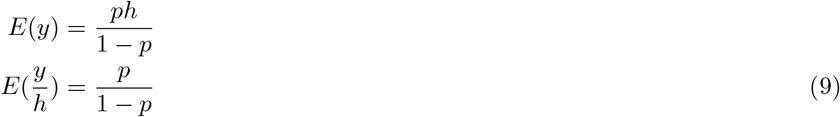

The link function in this parametrization of (7) is log-odds, i.e. 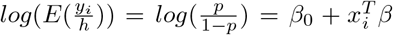. So, the likelihood for the Negative Binomial distribution ignoring the constant term is

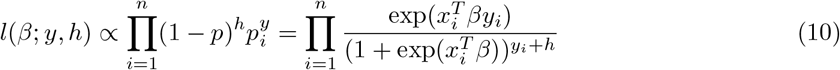

where 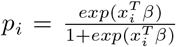. This *β* vector is assumed to include the intercept term, i.e. *β* = (*β*_0_, *β*_1_, …, *β*_*p*_).

### 2.3 Bayesian Negative Binomial Regression with Hierarchical Prior Structures (BNB)

The likelihood of NB (10) is not in a closed form and will require Polya-Gamma (PG) data augmentation. The auxillary variables will be sampled from Polya-gamma distribution with parameter *y*_*i*_ + *h* and 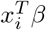. The Polya-Gamma distribution is the exact distribution needed to augment this posterior for simulation thus obtaining closed form posterior distributions that can be easily handled via Gibbs sampling. The method is useful when modeling proportions on the log-odds scale. Binary logistic regression [**?**] and negative binomial regression (NB) are the two fore frontiers that meets the criteria. To facilitate posterior sampling, we introduce a set of auxillary variables that follow Polya-Gamma distribution that are represented as scale mixtures of normals. Conditional on the latent variables, inference proceeds via straightforward Gibbs sampling. A Polya-Gamma variable, *w* ∼ *PG*(*b, c*) with *b >* 0 and *c* ∈ ℝ, can be defined as follows.

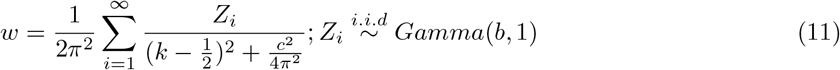

The variable distribution is similar to Gamma distribution and, as *b* increases, it becomes approximately normal [25]. So here 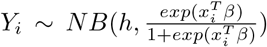 and 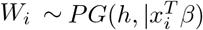. Similar, to logistic regression case, the joint posterior density of the parameter *β* and W with prior *π*(*β*) is obtained as

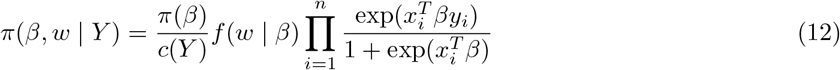

Here *c*(*Y*) is the marginal distribution of *Y*. We can see that

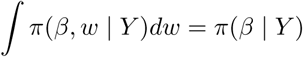

which is our targeted density. The conditional independence of *Y*_*i*_ and *W*_*i*_ implies that *π*(*w* | *β, y*) = *f* (*w* | *β*). Thus, we can draw from *π*(*w* | *β, y*) by making *n* independent draws from the Polya-Gamma distribution. The other conditional density *π*(*β* | *w, y*) is multivariate normal.

A hierarchical representation of the Horseshoe prior [9] is stated as

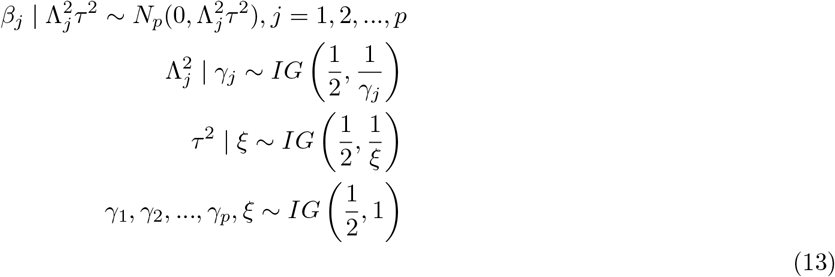

Here, Σ of the distribution of *β* is a diagonal matrix with elements 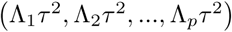. From equations (12), and the above hierarchical prior structure, the full posterior distribution is given by:

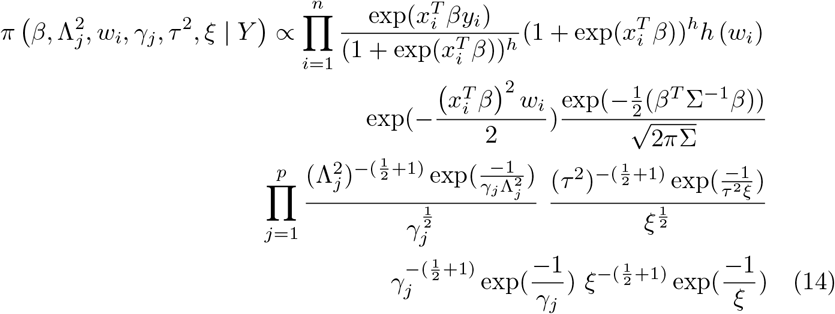

where *h*(*w*_*i*_) is obtained from

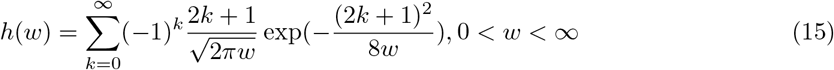

The conditional distributions required for our analysis follows:

The conditional density of *β* given *y, w* is

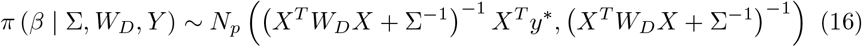

where, *W*_*D*_ and Σ are diagonal matrices where the elements are (*w*_1_, *w*_2_, …, *w*_*n*_), 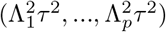respectively and, 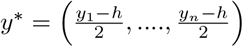.

The conditional density of *w*_*i*_ given *x*_*i*_, *β* is

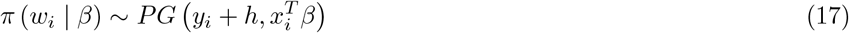

The conditional density of the hyper-parameters are as follows

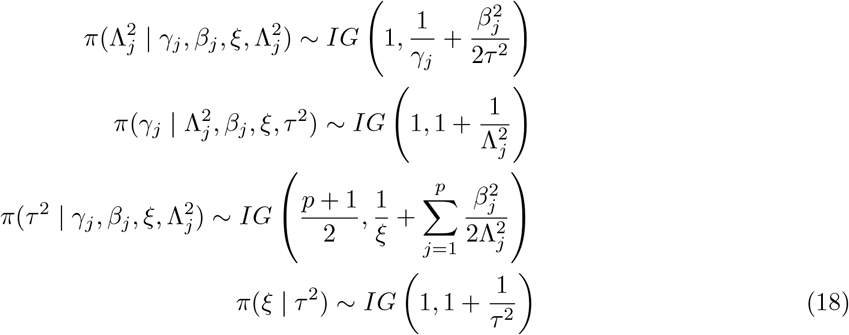

Again, here all the posterior densities are in the closed form, and follow simple densities like Normal, Polya-Gamma and Inverse-Gamma making sampling from them trivial. Exploiting the scale-mixture representation of the global-local shrinkage priors, it is straightforward to formulate the Gibbs sampler.

The hierarchical structure of the Dirichlet Laplace prior Bhattacharya:2015 is

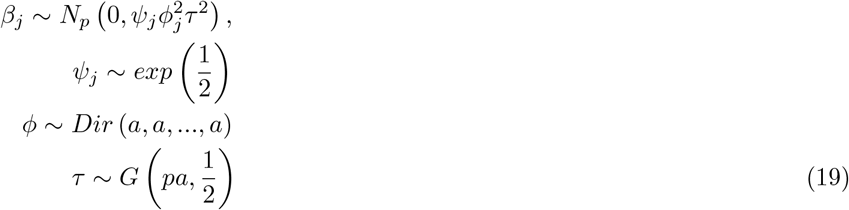

The conditional posterior distributions remain same for *β* | *y*_*i*_ and *w*_*i*_ | *β* is similar to that of equations (16) and (17).

The conditional density of the hyper-parameters as obtained similar to Theorem 2.2 in [14] are as follows:

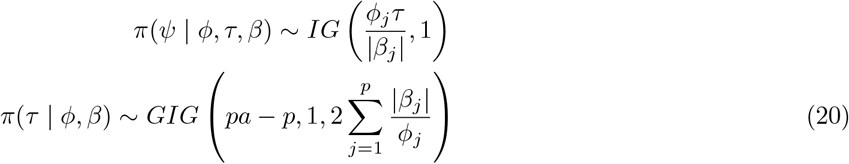

To sample *π*(*ϕ* | *β*_*j*_) sample 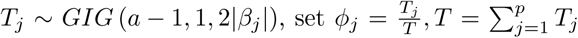. where *GIG*(*a, b, c*) is the Generalized Inverse Gaussian distribution with density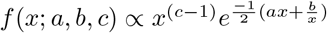.

The hierarchical structure of Double Pareto prior[26] is

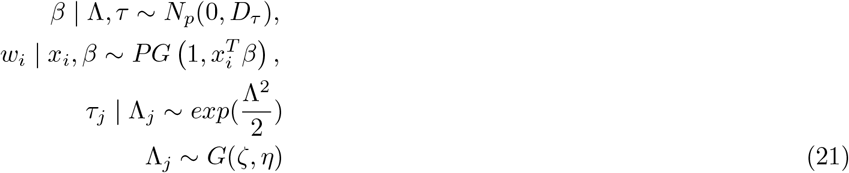

Again, the conditional densities of *β* | *y*_*i*_ and *w*_*i*_ | *β* remains same as (**??**). Here Σ = *D*_*τ*_ is a diagonal matrix with elements (*τ*_1_, *τ*_2_, …, *τ*_*p*_).

The conditional density of rest of the hyper-parameters are as follows:

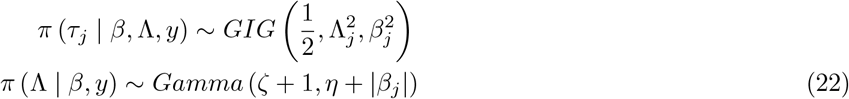

### 2.4 Bayesian Zero-Inflated Model with Hierarchical Prior Structures (BZINB)

Zero-inflated models represents the excess zeros, and a count distribution for the remaining values, thus forming a mixture of zeros. The model is very useful when there is an excess number of two types of zeros in the concerned response variable. By construction, zero-inflated models partition zeros into two types. The first type, typically referred to as a “structural” zero, corresponds to individuals who are not at risk for an event, and therefore have no opportunity for a positive count. The second type, termed the “at-risk” or “chance” zero, applies to a latent class of individuals who are at risk for an event but nevertheless have an observed response of zero[27]. For example, in our application with Covid-19 vaccine data set, examining the number of adverse events, the structural zeros might represent patients who had no adverse event thus have no recorded adverse event. In contrast, the at-risk zeros might correspond to patients with a single occurrence of adverse event which has been determined not clinically significant, thus contributes to at-risk zero. Similarly for RNA Seq datasets, where the genes are counts containing a high proportion of zeros, the zero-inflated models can be viewed as latent class models in which the classes are formed by the two types of zeros. The zero-inflated model has two parts that models consisting of negative binomial distribution, and the logit distribution.

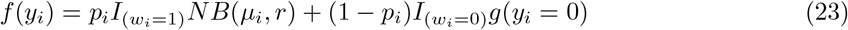

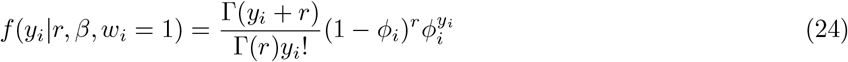

where 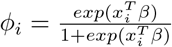 where *NB*(*µ*_*i*_, *r*) takes the form of equation 6. Here *y*_*i*_ is the count response for the ith individual. The latent indicator variable *wi* that takes values 1 and 0 with probabilities *p*_*i*_ and 1 − *pi* where *y*_*i*_ ∼ *NB*(*µ*_*i*_, *r*) and *y*_*i*_ = 0. The indicator*wi* variable is binary, thus modelled with logistic regression as follows:

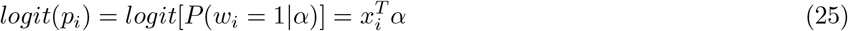

The negative binomial distribution has likelihood similar to equation 10 and the posterior distribution of the coefficients *β* are obtained from equation 12. With the hierarchical prior structures on the coefficients *β* from section 2.3, The posterior distribution for both *alpha* and *β* are modelled similarly as 2.3 citebhattacharyya2021applications, Neelon:2019.

## 3 Simulation

Here each of the simulation structure along with their parameters are defined.

### 3.1 BNB

The data is generated utilizing the Poisson-gamma mixture representation of NB as expressed in (4). In the data generation process, the true values of *β*, are defined. The data is generated as follows:

1. Calculate 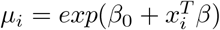
2. Get 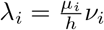, where *ν*_*i*_ ∼ *Gamma*(*h*, 1)
3. Generate *y*_*i*_ ∼ *Poi*(*λ*_*i*_)

The covariates are generated from multivariate normal distribution with mean vector 0 and covariance matrix Σ. 80% of the data set is reserved for the training set, and 20% for the test data set.

- S1: *n* = 200, *p* = 10 with a correlation of about *ρ* = 0.5 among the covariates. The coefficient vector is given by 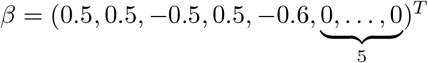 with 5 non-zero coefficients.
- S2: *n* = 120, *p* = 10, *ρ* = 0.2 Five non-zero coefficients. 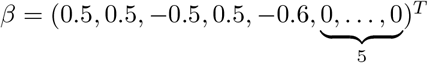
- S3: *n* = 50, *p* = 10, coefficients similar to S2.
- 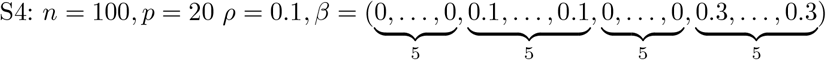
- 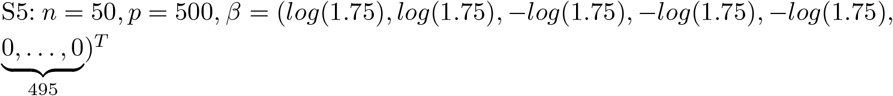

Here we have considered two broad settings of design matrices comprising *p* = 200 and *p* = 500 covariates. The sampling of *β* for *p > n* with *p* = 500 is conducted from a Gaussian distribution and follows the fast sampling algorithm of [28]. For prediction accuracy, the mean squared error (MSE) is used as a prediction accuracy criteria, which is defined as 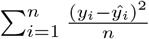. In terms of variable selection performance, the number of the truly nonzero coefficients which are incorrectly set to zero (FP), and the number of the true zero coefficients which are correctly set to zero (FN). The higher the values of FP, and the lower the values of FN, the better the variable selection performance is. The variable selection is determined by posterior credible intervals. For each simulation setting, 100 datasets were generated, and MSE, results were calculated by averaging over these 100 datasets. MCMC is used for sampling which relies on block-updating the sets of parameters. The no. of simulation runs is 10000 with 6000 as burn-in. Trace plots, auto-correlation plots, Geweke z-statistics [29] were some of the diagonistic criterion. R pacakge coda [30] was used to conduct the posterior sample diagnosis and pgdraw [31] [32] was used to generate the Polya-Gamma random variates. All the computations have been carried out in RStudio. The R package coda [30] was used to conduct the posterior sample diagnosis and pgdraw [31] [32] was used to generate the Polya-Gamma random variates.

### 3.2 BZINB

The following simulation scenarios were set up to understand the behaviour of the model. We followed similar data generation process as of [27].

- Sim1: *n* = 1000, *p* = 4, *α* = (0.5, 0.5, −0.25, 0.25), *β* = (0.5, −1.00, 0.75, −0.25), *r* = 1
- Sim2: *n* = 500, *p* = 10, *α* = (0.5, 0.5, −0.25, 0.25, 0.5, 0.5, −0.25, 0.5, 0.5, −0.25),*β* = (0.5, 0.5, 0.75, −0.25, 0.5, 0.75, 0.25, 0.75, −0.25), *r* = 1
- Sim3: 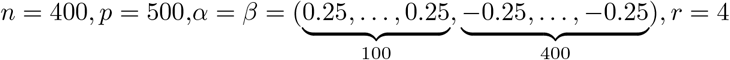

## 4 Simulation Results

Table 1 shows the variable selection performance and MSE along with their standard deviations for these simulation scenarios for BNB model. Comparing S1, S2, and S3 accuracy and sensitivity were similar with the decrease in sample size for all the three priors. The MSE for *β* and their standard errors increases for all the three priors as the sample size decreases. From S1 and S2 and S3 are three scenarios having similar settings with the N/P ratio changing, there is a clear trend that with the decrease in sample size increases the MSE, S1 and S2 has 100% sensitivity implying all the non-zero *β* can be identified. There is not much of an impact of correlation on the results. S5 has 1% non-zero *β* with low effect sizes but almost 67% of the time they are identified by the 2 priors DL and Horseshoe. DP fails to identify the non-zeros, though keeping the MSEs low.

**Table 1:** Variable Selection Performance among BNB Simulation Scenarios

| Simulation Scenarios |  |  |  |  |  |
| --- | --- | --- | --- | --- | --- |
| Priors | s1 | s2 | s3 | s4 | s5 |
| N,P | 200,10 | 120,10 | 50,10 | 100,20 | 50,500 |
| Accuracy |  |  |  |  |  |
| Horseshoe | 0.993(0.026) | 0.996(0.020) | 0.958(0.068) | 0.702(0.048) | 0.827(0.066) |
| Dirichlet Laplace | 0.991(0.029) | 0.989(0.035) | 0.962(0.065) | 0.722(0.053) | 0.829(0.064) |
| Double Pareto | 0.992(0.027) | 0.997(0.017) | 0.956(0.073) | 0.702(0.046) | 0.832(0.071) |
| Sensitivity |  |  |  |  |  |
| Horseshoe | 1.000(0.000) | 1.000(0.000) | 0.936(0.110) | 0.514(0.088) | 0.662(0.123) |
| Dirichlet Laplace | 1.000(0.000) | 1.000(0.000) | 0.950(0.096) | 0.563(0.094) | 0.668(0.114) |
| Double Pareto | 1.000(0.000) | 1.000(0.000) | 0.932(0.114) | 0.516(0.084) | 0.012(0.003) |
| Specificity |  |  |  |  |  |
| Horseshoe | 0.986(0.051) | 0.992(0.039) | 0.980(0.072) | 0.889(0.031) | 0.994(0.005) |
| Dirichlet Laplace | 0.982(0.058) | 0.978(0.069) | 0.989(0.031) | 0.880(0.045) | 0.977(0.003) |
| Double Pareto | 0.984(0.055) | 0.994(0.034) | 0.988(0.033) | 0.887(0.034) | 0.999(0.005) |
| MSE |  |  |  |  |  |
| Horseshoe | 0.194(0.006) | 0.201(0.022) | 0.229(0.040) | 0.101(0.010) | 0.003(0.000) |
| Dirichlet Laplace | 0.195(0.014) | 0.201(0.022) | 0.228(0.039) | 0.103(0.010) | 0.005(0.000) |
| Double Pareto | 0.194(0.014) | 0.201(0.022) | 0.229(0.039) | 0.102(0.011) | 0.003(0.000) |

In summary, it is obvious that the simulation results has demonstrated the use of these three priors unanimously with the low standard errors across datasets. Trace plots, Geweke diagnostics and Monte Carlo standard errors were indicative of convergence and showed reasonable mixing fora range of model parameters. We tested the sampler for various settings of *n, p* and coefficients. Comparing S2, and S3 the MSE for *β* and their standard errors increases for all the three priors and drastically reduces the MSE of the fitted values as well. The increase in correlation in S4 than S3 doesn’t seem have much effect. As *nandp* increases from S1 to S2 there is a substantial decrease in MSE of *β* but not for the fitted values. S5, S7 and S12 compare different sample size under similar coefficient and simulation settings. MSE and their standard error decreases as their sample size increases. S9, S10 and S11 again compare sample sizes *n* = 200, 100, 8 for a different set of coefficient settings. Here all the *β*s were non-zero. Again the decrease in MSE with the increase of sample size is noticeable and also in the MSE of fitted values. In the all the above *n > p* scenarios we see that Horseshoe, and Double Pareto perform comparatively better than Dirichlet Laplace. For *p > n* scenarios S13 and S6, with the increase in the *n/p* ratio increases MSE increases. In summary, it is obvious that the simulation results has demonstrated the use of these three priors unanimously. We build on the R codes from this package and extend it with the three different prior structures.

In BZINB model, Horseshoe prior clearly outperforms the other two priors across simulation scenarios. The Sim3 *p > n* case has similar performance across priors. The variable selection performances are also measured with the MSEs and all the three priors performed reasonably Table 2.

**Table 2:**
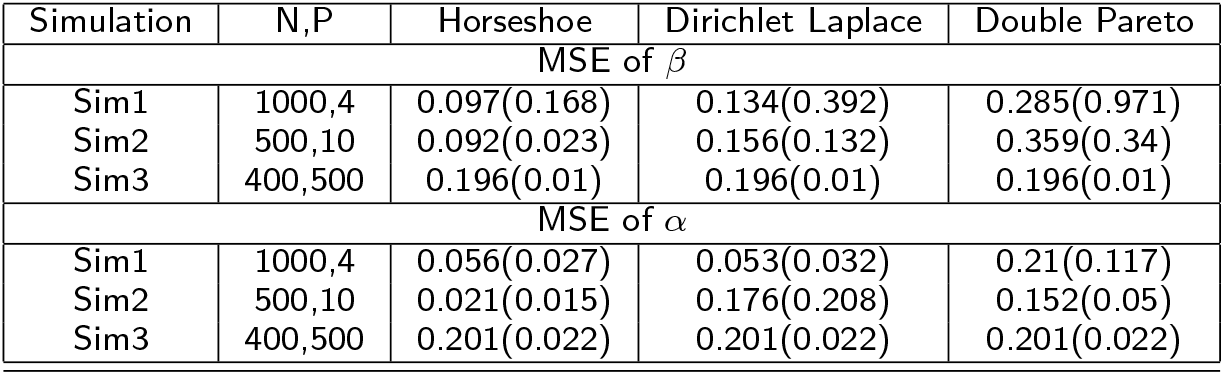
Comparison of MSE among BZINB Simulation Scenarios

| Simulation | N,P | Horseshoe | Dirichlet Laplace | Double Pareto |
| --- | --- | --- | --- | --- |
| MSE of $\beta$ | | | | |
| Sim1 | 1000,4 | 0.097(0.168) | 0.134(0.392) | 0.285(0.971) |
| Sim2 | 500,10 | 0.092(0.023) | 0.156(0.132) | 0.359(0.34) |
| Sim3 | 400,500 | 0.196(0.01) | 0.196(0.01) | 0.196(0.01) |
| MSE of $\alpha$ | | | | |
| Sim1 | 1000,4 | 0.056(0.027) | 0.053(0.032) | 0.21(0.117) |
| Sim2 | 500,10 | 0.021(0.015) | 0.176(0.208) | 0.152(0.05) |
| Sim3 | 400,500 | 0.201(0.022) | 0.201(0.022) | 0.201(0.022) |

## 5 Real Data Application

Here three real data applications are considered. In all the applications the count data and the excess amounts of zeros in the outcome made BNB and BZINB regression a best fit for the data. The variables selected by the three priors for BNB and BZINB, Poisson regression and NB models for each of the datasets is in the table 3. For the three priors, the starting values are chosen as the maximum likelihood estimates from NB Regression. The MSE for the coefficients for the tree priors are obtained by 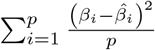, *p* is the number of coefficients, *β*_*i*_: estimates obtained from NB regression and 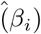 are the posterior mean estimates. This MSE measure can also be interpreted as a departure from the frequentist estimates.

**Table 3:** Variable Selection in Real World Data

| Datasets |  |  |  |
| --- | --- | --- | --- |
| Methods | Selected Variables | MSE.$\beta$ | MSE.Y |
| **Biochemists** |  |  |  |
| NB with Horseshoe | $x_3, x_5$ | 0.017 | 4.19 |
| NB with Dirichlet Laplace | $x_3, x_5$ | 0.016 | 4.19 |
| NB with Double Pareto | $x_1, x_3, x_5$ | 0.015 | 4.18 |
| ZINB with Horseshoe | $x_3, x_5$ | 0.011 | 8.07 |
| ZINB with Dirichlet Laplace | $x_3, x_5$ | 0.01 | 7.84 |
| ZINB with Double Pareto | $x_3, x_5$ | 0.01 | 8.72 |
| Poisson | $x_1, x_2, x_3, x_5$ | 0 | 3.75 |
| Negative Binomial | $x_1, x_3, x_5$ | 0 | 3.79 |
| Zero-Inflated Poisson | $x_1, x_2, x_3, x_5$ | 0 | 9.17 |
| Zero-Inflated Negative Binomial | $x_1, x_2, x_5$ | 0 | 8.26 |
| **Nuts** |  |  |  |
| NB with Horseshoe | $x_5, x_6, x_7$ | 0.309 | 607.5 |
| NB with Dirichlet Laplace | $x_5, x_6, x_7$ | 0.312 | 608.4 |
| NB with Double Pareto | $x_5, x_6, x_7$ | 0.312 | 605.9 |
| Poisson | $x_5, x_6, x_7$ | 0 | 184.05 |
| Negative Binomial | $x_7$ | 0 | 176.23 |
| **NMES** |  |  |  |
| NB with Horseshoe | $x_1, \dots, x_6$ | 0.004 | 64.40 |
| NB with Dirichlet Laplace | $x_1, \dots, x_6$ | 0.004 | 64.38 |
| NB with Double Pareto | $x_1, \dots, x_6$ | 0.004 | 64.39 |
| Poisson | $x_1, \dots, x_12, x_14, \dots, x_16$ | 0 | 47.31 |
| Negative Binomial | $x_1, \dots, x_6, x_8, \dots$ | 0 | 59.26 |
| **Covid-19 vaccine** |  |  |  |
| NB with Horseshoe | $x_1, x_2, x_3, x_4, x_6, x_7, x_{10}, \dots, x_{19}$ | 6.21 | 390.36 |
| NB with Dirichlet Laplace | $x_2, x_4, x_6, \dots, x_{12}, x_{15}, x_{16}, x_{19}$ | 6.46 | 390.24 |
| NB with Double Pareto | $x_7, x_9, x_{10}, x_{13}, x_{14}, x_{16}$ | 5.14 | 390.27 |
| ZINB with Horseshoe | $x_1, \dots, x_5, x_7, x_8, x_9, x_{11}, \dots, x_{15}, x_{17}, x_{18}, x_{20}$ | 6.06 | 427.11 |
| ZINB with Dirichlet Laplace | $x_7, x_9, x_{10}, x_{13}, x_{14}, x_{16}$ | 73.5 | 386.72 |
| ZINB with Double Pareto | $x_1, \dots, x_{20}$ | 5.88 | 407.19 |
| Poisson | $x_1, x_2, x_4, x_5, x_7, \dots, x_{14}, x_{16}, x_{17}, x_{18}, x_{20}$ | 2.24 | 1566.97 |
| Negative Binomial | $x_1, \dots, x_5, x_7, \dots, x_{10}, x_{11}, x_{13}, \dots, x_{15}, x_{17}, \dots, x_{20}$ | 2.42 | 1566.97 |
| Zero-Inflated | $x_1, x_2, x_5, x_8, x_9, x_{10}, x_{11}, x_{13}, x_{14}, x_{16}, x_{17}, x_{18}, x_{20}$ | 3.5 | 1546.08 |
| **PBMC RNA-Seq** |  |  |  |
| NB with Horseshoe | 2742, 6369, 9338, 3665, 7203, 11458, 2217, 4360, 4368 | 0.00 | 20.35 |
| NB with Dirichlet Laplace | 2742, 13706, 6369, 9338, 11458, 3665, 7203, 2217, 4360, 4368 | 0.00 | 19.92 |
| NB with Double Pareto | 2742, 13706, 6369, 9338, 11458, 7203, 2217, 3665, 4368, 11191 | 0.00 | 20.40 |
| ZINB with Horseshoe | 74, 267, 1521, 1804, 2612, 3417, 3695, 6049, 8146, 8285 | 0.00 | 20.43 |
| ZINB with Dirichlet Laplace | 74, 153, 267, 904, 1804, 3417, 6049, 6680, 7526, 8146 | 0.00 | 7.48 |
| ZINB with Double Pareto | 74, 267, 1804, 1983, 2323, 3124, 3317, 3417, 5528, 6049 | 0.00 | 6.06 |
| **Covid-19 RNA-Seq** |  |  |  |
| NB with Horseshoe | 4876, 4903, 7767, 8998, 9016, 9856, 10183, 11222, 11598, 12202; | 0.0 | 1016.338 |
| NB with Dirichlet Laplace | 4903, 4995, 7767, 7798, 8523, 10416, 11222, 12202, 13109, 16202; | 0.0 | 3276.73 |
| NB with Double Pareto | 8523, 8998, 9016, 9301, 9856, 12202, 13109, 14012, 14476, 15265; | 0.0 | 4971.96 |
| ZINB with Horseshoe | 22, 58, 202, 231, 272, 280, 297, 314, 404, 523; | 0.0 | 7.11 |
| ZINB with Dirichlet Laplace | 193, 219, 293, 391, 487, 514, 589, 625, 697, 724; | 0.0 | 1.99 |
| ZINB with Double Pareto | 76, 113, 163, 179, 202, 279, 356, 366, 428, 446; | 0.0 | 3.97 |

### 5.1 No. of PhD publications

The first one deals with the number of publications produced by Ph.D. biochemists of [33]. It is available in the R package “pscl”[34].The response variable is the number of articles in the last three years of Ph.D. Five explanatory variables were used. They are: the gender (*x*_1_), the marital status (*x*_2_), the number of children under age six (*x*_3_), prestige of Ph.D. program (*x*_4_), and the number of articles by the mentor in last three years (*x*_5_). In this application, the response variable is following NB distribution. (*x*_3_) and (*x*_5_) were selected by the Bayesian methods. The NB, poisson followed by BNB methods were suitable for the data set, as per the MSE of fitted values.

### 5.2 Nuts data

In the second real data application, we considered the nuts dataset [18]. Here, *n* = 52 and *p* = 7. The nuts dataset defines the squirrel behavior and several features of the forest across different plots in Scotland’s Abernathy Forest. It is available in the R package “COUNT”[35].The response variable is the number of cones stripped follows negative binomial distribution. The explanatory variables are: the number of trees per plot (*x*_1_), the number of DBH per plot (*x*_2_), mean tree height per plot (*x*_3_), canopy closure (as a percentage) (*x*_4_), standardized number of trees per plot (*x*_5_), standardized mean tree height per plot (*x*_6_), standardized canopy closure (as a percentage) (*x*_7_). Here we use *x*_5_, *x*_6_, *x*_7_.

The variables chosen by all the methods except NB were *x*_5_, *x*_6_, *x*_7_. So all the models seem reasonable except NB. The variables selected in the first two datasets (Biochemists, and NUTS) are included withing the set of the variables selected either by the traditional Poisson or NB regression.

### 5.3 US National Medical Expenditure Survey

The third data set originated from the US National Medical Expenditure Survey (NMES) conducted in 1987 and 1988. The NMES is based upon a sample of the civilian non-institutionalized population and individuals admitted to long-term care facilities during 1987. The data are a sub-sample of individuals ages 66 and over all of whom are covered by Medicare (a public insurance program providing substantial protection against health-care costs). It is available in the R package “AER”[36]. It is a data frame containing 4,406 observations on 19 variables. The response variable considered is the number of physician office visits among other type of count variables present (emergency visits, no. of non-physician hospital outpatient visits etc.). We have considered 14 dependent variables hospital: number of hospital stays *x*_1_; health:self-perceived health status *x*_2_, levels are “poor”, “average”, “excellent”; chronic:number of chronic conditions *x*_3_; adl: indicator of whether individual having limits in activities of daily living *x*_4_ levels: “limited”,”normal”; region: indication region of the individual *x*_5_ levels northeast, midwest, west, other; age *x*_6_; afam (race): If African-American *x*_7_; gender:male/female *x*_8_; married: marital status *x*_9_; school:number of years of education *x*_10_; income (USD) *x*_11_; employment *x*_12_; insurance: whether the individual is covered by private insurance yes/No *x*_13_; medicaid *x*_14_. The MSE for the fitted values are also given. Figure 1 the posterior distribution of the 14 variables and their respective confidence intervals. For the NMES data, the three priors selects 10 important out of 14 variables, Poisson selects 13 and NB 8 of them. It seems that the three priors perform better than NB where they do include the relevant variables such as limited activity level *x*_4_ and race *x*_7_ but doesn’t do over-fitting such as Poisson.

**Figure 1:**
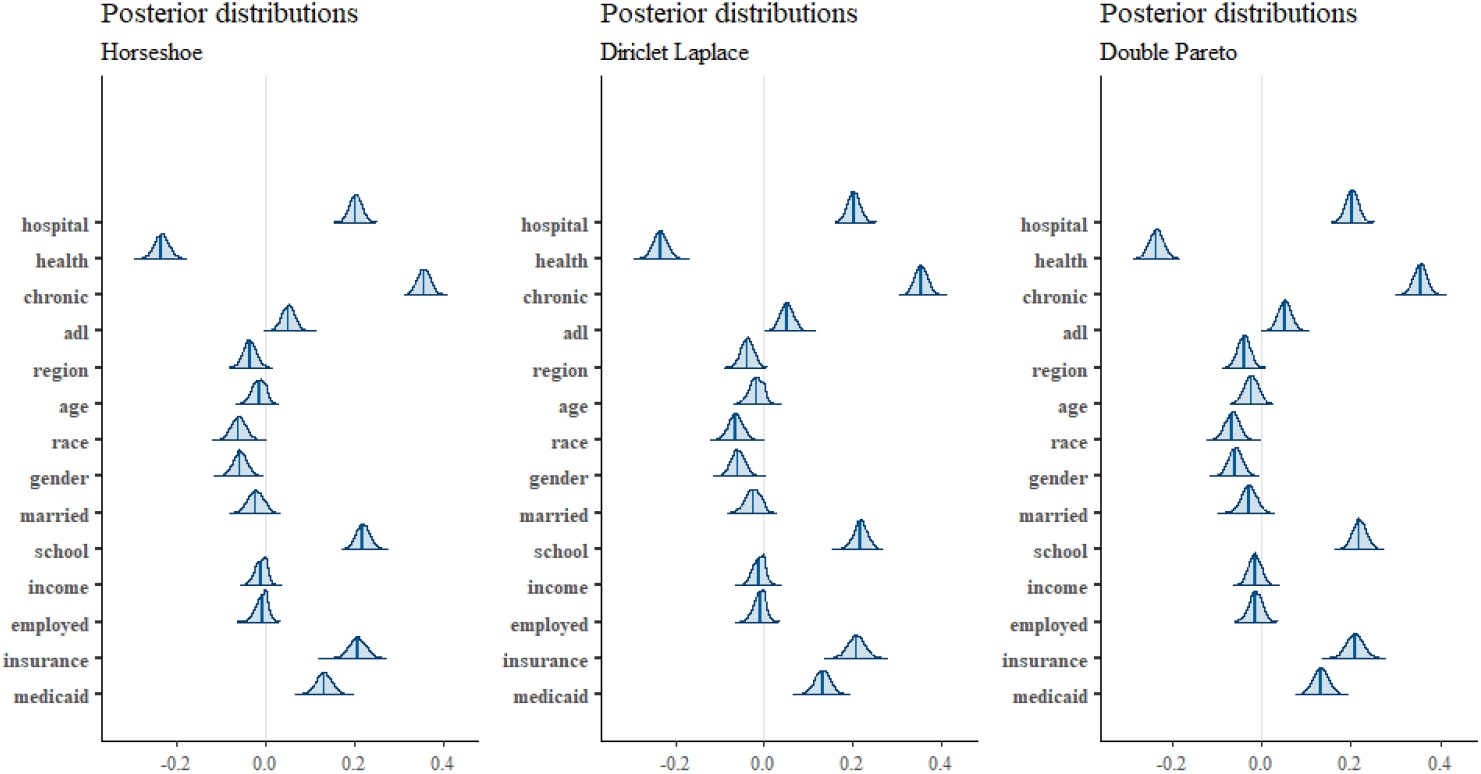
Posterior distribution with 95% credible intervals: NMES Data

### 5.4 PBMC RNA-Seq data

The data set that is analyzed here is taken from https://satijalab.org/seurat/articles/pbmc3ktutorial.html. The RNASeq data is analyzed as a per gene model where each gene is the outcome and the cell type as the covariate. The top 10 genes selected by BNB and BZINB are in Table 3. The genes common between H and DL are 2742, 6369, 9338, 3665,7203,11458, 2217, 4360,4368.The genes common between DL and DP are 2742,13706,6369,9338,11458,7203, 2217, and 4368. The genes common between DP and H are 2742,6369,9338,3665,11458,7203,2217,3665,4368,4368. The genes common between three priors are 2742, 6369, 9338,7203,11458,3665, 2217, 4368. The common genes between H and DL are 74, 267, 1804,3417,6049,8146. The genes that belong to the intersection of DL and DP are 74, 267,1804, 3417, and 6049. The genes common between DP and H are 74, 267, 1804, 3417, and 6049. The genes common between the three priors are 74, 267, 1804, 3417, and 6049. The genes selected by both the models are selected by ranking them by the least MSE.

### 5.5 Covid-19 Vaccine Data

This data set consisted of the Covid-19 vaccine administered over the year 2021 and the symptoms (adverse events) gathered from the administration of vaccines. This data is a part of the Vaccine Adverse Event Reporting System (VAERS) which was created by the Food and Drug Administration (FDA) and Centers for Disease Control and Prevention (CDC) to receive reports about adverse events that may be associated with vaccines.The variables that are considered are age (*x*_1_), sex (*x*_2_), if there is life threat or not (*x*_3_), if there was emergency room visit (*x*_4_), if hospitalized (*x*_5_), number of hospital days (*x*_6_), no. of extended stay (*x*_7_), disability status (*x*_8_), recovery status (*x*_9_), medical history (*x*_10_), other medications (*x*_11_), laboratory data (*x*_12_), disease during vaccination (*x*_13_), prior vaccination status (*x*_14_), allergy status (*x*_15_), doctor’s office visit (*x*_16_), emergency visit (*x*_17_), vaccination route (*x*_18_), vaccination dose (*x*_19_), vaccine manufacturer (*x*_20_).The dimension of the cleaned data set post-processing was having 100 samples and 20 variables. The number of days between vaccination and onset of adverse symptoms is treated as the response variable (*y*) that had a median of 1 day, mean of 7 days, maximum minimum of 0, and 2.43 years respectively. The results from the BNB model, BZINB model, and both these models with respective priors are given in Table 3. We applied the region of practical equivalence (ROPE) method [37] was utilized to select the variables after obtaining the posterior samples. The variables that were significant by the ROPE method with 5% cut-off for ROPE by mainly the Horseshoe and the DP priors for both the BNB and BZINB models belonged to the super set of the set of the variables. The variables age (*x*_1_), sex (*x*_2_), if there is life threat or not (*x*_3_), if there was emergency room visit (*x*_4_), no. of extended stay (*x*_7_), other medications (*x*_11_), laboratory data (*x*_12_), disease during vaccination (*x*_13_), prior vaccination status (*x*_14_), allergy status (*x*_15_) belonged to the intersection of the two sets of variables identified by the BNB and BZINB models with Horseshoe prior, which was a significant overlap along with matching the variables identified by the traditional methods without shrinkage priors. In general, the BZINB with DP and Horseshoe priors were able to select more variables than the BNB model. Specifically, variables such as medical history (*x*_10_), prior vaccination status (*x*_14_), vaccination route (*x*_18_), vaccine manufacturer (*x*_20_) were some of the interesting features that seem to influence the results. All the shrinkage prior models surpassed the traditional models in their MSE of fitted values.

### 5.6 Covid-19 RNASeq Data

The data is taken from the article [38]. We selected the GSE152075 dataset from Gene Expression Omnibus (GEO: https://www.ncbi.nlm.nih.gov/gds), which contained RNA-seq data from 430 SARS-CoV-2 positive and 54 negative patients [**?**]. The data is analyzed similarly with the help of per gene model where each RNA-seq (count variable) is modelled against the covid-19 positivity status which is a binary variable. Upon pre-processing the following top 10 genes were selected by ranking the genes with the lowest MSE with the BZINB and BNB methods are in Table 3. The BZINB method was able to select about 30 genes that were not selected by the top BNB method. With the BNB method,there were 4 genes commonly selected by DL and H which are 4903, 7767, 11222, and 12202.The common genes between DL and DP are 8523,12202,13109 and between DP and H are 8998, 9016, 9856, 12202. Gene no. 12202 was selected by all three priors with BNB model.

## 6 Discussion

Our simulations showed that the approach performs well across a range of scenarios. The numerical results provide additional numerical and theoretical insights into the properties of global-local shrinkage priors including high-dimension case. Variable selection is a very helpful procedure for improving computational speed and prediction accuracy by identifying the most important variables that related to the response variable. The number of counts for each observation if large can make the sampler perform poorly as the Polya-Gamma augmentation will not be efficient as it require generation of Polya-Gamma random variates equal to the number of observations[16]. The generation of such random variates is also time consuming.The Polya-Gamma procedure is in general fast, easy to implement and flexible. The rate at which one can generate Polya-Gamma random variates is a key factor in the efficiency of the Polya-Gamma scheme; hence, building fast samplers is essential. R packages such as “bayesreg”[31] deals with high-dimensional Bayesian regularised regression. Alternative models such as the BNB model, BZINB models [27], truncated models, or quantile count models provide potential future research guidelines. The use of shrinkage priors with next generation sequencing data such as RNA-Seq data with Zero-Inflated or Negative Binomial models are also areas that needs further exploration [2]. All the three priors have their own advantage and caveats. A computationally efficient Bayesian approach for variable selection is proposed here that performs quite well in simulation scenarios and provide consistent results in these different case studies. One disadvantage of data augmentation schemes is that the number of latent variables is of the order of sample size. Hence for large *n*, the computation can slow down, as we have seen in settings with sample sizes greater than 1000. However, this disadvantage is offset by the gains we have over the traditionally used Metropolis-Hastings, which requires choosing proposal distributions and would probably generate a considerable number of rejection steps. Additionally, the model can be extended with other shrinkage priors, and other models such as hurdle models. More generally, the proposed method can be applied in scenarios where interest lies in modeling count data within a Bayesian inferential framework and exhaustive comparison of existing shrinkage priors in the literature. We believe that the rigorous, yet simple and systematic nature of Bayesian inference coupled with the latest advances in technology in high dimensional and next generation sequencing with RNA Seq data might strongly help in contributing to expanding the research field.

## Data Availability

The datasets used and/or analysed are publicly available and information about it is included in this article.

## 7 Acknowledgements

This work was supported by the National Institute of Health grant P42 ES023716 to principal investigator: Dr S Srivastava and the National Institute of Health grant 1P20 GM113226 to principal investigator: Dr C McClain. Dr. Rai was also partially supported by Wendell Cherry Chair in Clinical Trial Research.

## Abbreviations

SSVS: Stochastic Search Variable Selection
GL: global-local
HS: Horseshoe
DL: Dirichlet Laplace
DP: Double Pareto
PG: Polya-Gamma
DA: Data-Augmentation
MCMC: Markov Chain Monte Carlo
MSE: Mean Squared Error
VS: variable selection
BZINB: Bayesian Zero-Inflated Negative Binomial
BNB: Bayesian Negative Binomial
NB: Negative Binomial
ZINB: Zero-Inflated Negative Binomial;

## Availability of data and materials

The datasets used and/or analysed are publicly available and information about it is included in this article. Not applicable

## Consent for publication

Not applicable

## Authors’ contributions

A.B. has contributed to the methodology, data collection, analysis, and writing of the manuscript. R.M. and S.P. has contributed to the methodology. S.N.R. has contributed to developing ideas, analysis and valuable comments. All authors have contributed to the final preparation of the manuscript.

